# The Montreal Cognitive Assessment: Normative Data from a Large, Population-Based Sample of Healthy Adults in China

**DOI:** 10.1101/2023.12.18.23300135

**Authors:** Qiang Wei, Baogen Du, Yuanyuan Liu, Shanshan Cao, Shanshan Yin, Ying Zhang, Tongjian Bai, Xingqi Wu, Yanghua Tian, Panpan Hu, Kai Wang

**Affiliations:** Department of Neurology, the First Affiliated Hospital of Anhui Medical University, Hefei, Anhui 230022, China; Anhui Province Key Laboratory of Cognition and Neuropsychiatric Disorders, Hefei, Anhui 230032, China; Collaborative Innovation Center of Neuropsychiatric Disorders and Mental Health, Anhui Province, Hefei, Anhui 230032, China; Wuhan WuDong Hospital, Wuhan, Hubei 430084, China; Department of Psychology and Sleep Medicine, The Second Affiliated Hospital of Anhui Medical University, Hefei, Anhui 230601, China; Institute of Artificial Intelligence, Hefei Comprehensive National Science Center, Hefei, Anhui 230088, China; The School of Mental Health and Psychological Sciences, Anhui Medical University, Hefei, Anhui 230032, China

**Keywords:** Montreal Cognitive Assessment, Healthy adults, Normative data, Regression analyses

## Abstract

**Background:** The Montreal Cognitive Assessment (MoCA) is a valuable tool for detecting cognitive impairment, but its accuracy is significantly influenced by demographic and socio-cultural factors. Consequently, the development of appropriate normative values becomes particularly crucial in ensuring its reliable use and interpretation.

**Objective:** Generate MoCA normative values based on demographics for healthy chinese adults.

**Methods:** The assessment conducted in this study utilizes the MoCA scale, specifically employing the Mandarin-8.1 version (Chinese Mandarin version). Based on the geographical distribution of administrative regions in mainland china, this study recruited a total of 3,097 healthy individuals aged over 20 years. Drawing on insights from prior normative studies, we performed multiple linear regression analysis, incorporating age, gender, and education level as predictor variables, to examine their associations with the total score and sub-cognitive domains of MoCA. Subsequently, we established normative values and cut-off values stratified by age and education level.

**Results:** The participants in this study (n=3,097) exhibit a balanced gender distribution, with an average age of 54.46 years (SD 14.38) and an average education period of 9.49 years (SD 4.61). The study population demonstrates an average MoCA score of 23.25 points (SD 4.82). The findings from the multiple linear regression analysis indicate that the total score of MoCA is influenced by age and education level, collectively accounting for 46.8% of the total variance. Higher age and lower education level are correlated with lower MoCA total scores.

**Conclusion:** This study offers normative MoCA values specific to the Chinese population. Furthermore, the research findings indicate that a score of 26 may not represent the most optimal cut-off value. When assessing MoCA scores, it is imperative to comprehensively account for the participants’ age and educational background.

## Introduction

Age-related cognitive impairment represents a significant public health and social challenge in the contemporary era. In 2018 alone, approximately 50 million individuals worldwide were affected by this condition, with projections estimating that the number will triple by 2050, leading to a staggering economic loss of nearly $4 trillion.^[1]^ For cognitive disorders, early identification is of paramount importance to maximize the effectiveness of interventions, which may include counseling, psychoeducation, cognitive training, and medication.^[2]^ Owing to the absence of effective treatments for advanced dementia, the significance of early diagnosis and intervention during the mild cognitive impairment (MCI) stage has gained widespread recognition as a pivotal approach in disease management. These strategies hold the potential to impact long-term outcomes.^[3]^ The early detection and identification of cognitive decline necessitates a straightforward, easily comprehensible, and highly diagnostic tool. Historically, the Mini-Mental Status Examination (MMSE) served as the widely adopted assessment tool.^[4]^ Nevertheless, owing to the absence of executive function assessment,^[5]^ the MMSE demonstrates limited sensitivity in identifying mild cognitive impairment.^[6, 7]^ Subsequently, the Montreal Cognitive Assessment (MoCA) emerged as a viable alternative,^[8]^ a widely utilized international neuropsychological screening tool, proficient in evaluating a subject’s global cognitive function. In comparison to the MMSE, MoCA incorporates additional tasks to assess visuospatial abilities, as well as specific evaluations of frontal/executive function and attention.^[9]^ Consequently, it exhibits heightened sensitivity and specificity in detecting cognitive impairment, particularly mild cognitive impairment.^[10, 11]^ And it has been widely recognized and widely used in clinical and scientific research. The MoCA scale employs cognitive tasks that are rapid, sensitive, and easily manageable, and it has undergone multiple revisions throughout its usage. Over an extended period of MoCA implementation, scores below 26 out of 30 were deemed indicative of cognitive impairment; however, empirical investigations have established that despite its high sensitivity, this threshold exhibits limited specificity.^[12, 13]^

Over the past decade, the scale has undergone translation into multiple languages and validation in diverse populations encompassing various ages and educational backgrounds.^[13-32]^ However, owing to variations in cultural and linguistic practices across different countries and regions, the performance of MoCA may exhibit variability. As a result, it is particularly important to make reasonable localization revisions to the MoCA scale and develop appropriate local normative values. Currently, this approach has been implemented in numerous countries.^[15, 16, 20-26, 28-31, 33]^

Nevertheless, as far as our knowledge extends, there exists a deficiency in normative values specifically tailored to the Chinese population and its cultural attributes. This dearth might curtail the attention dedicated to the societal concern of cognitive impairment and impede the implementation of appropriate early interventions. To address this gap, this study used Mandarin-8.1 version (Chinese Mandarin version), which was localized and revised based on the language and cultural characteristics of the Chinese population. The primary objective was to furnish normative MoCA data stratified by age and education, derived from a large, geographically diverse, population-based sample.

## Methods

### Ethical approval

All participants provided written informed consent, and the research scheme was approved by the Anhui Medical University Ethics Committee.

### Study design and participants

This study comprised a total of 3,097 cognitively healthy individuals from various provinces, autonomous regions, and municipalities directly under the Central Government of Mainland China. Participants who meet the following eligibility criteria can undergo MoCA assessment.

All participants met the following inclusion criteria: 1) age of 20 years or older, 2) proficiency in speaking and understanding Chinese, 3) provision of informed consent, 4) capability to effectively complete the MoCA test, 5) ability to take care of themselves in daily life, and 6) no complaints of cognitive impairment. Exclusion criteria encompassed: 1) presence of dementia, mild cognitive impairment, or any known cognitive impairment attributed to neurological or non-neurological disorders or treatments (e.g., chemotherapy), 2) history of stroke within the past five years, 3) manifestation of moderate or severe mental disorders, and 4) presence of other conditions precluding successful completion of the MoCA test.

### Measurements

Compared with the original English version of MoCA 8.1, the Mandarin-8.1 version has been localized and revised based on the language characteristics and cultural habits of the Chinese population, mainly manifested in the vocabulary used in alternating connection tests, memory tests, attention tests, sentence repetition tests, and word fluency tests. The total score comprises 30 points, and the assessment can be completed in approximately 10 minutes. The scoring is divided into various cognitive domains, including visual space/executive function (0-5 points), naming function (0-3 points), attention function (0-6 points), language function (0-3 points), abstraction function (0-2 points), delayed recall function (0-5 points), and orientation function (0-6 points). All recruiters and evaluators have received training from neurologists well-versed in Neuropsychology testing methodologies.

### Statistical analysis

This study used SPSS 20.0 for statistical analysis, and multiple linear regression methods were used to analyze the relationship between age, gender, education level, MoCA total score, and various sub cognitive domain scores. To obtain reliable normative values, age was categorized into six groups (<40 years old, 40-49 years old, 50-59 years old, 60-69 years old, 70-79 years old, and ≥80 years old), while education level was classified into four groups (illiteracy, primary school, middle school, and high school and above). Moreover, the research results were used to determine different cut-off values based on the obtained means and standard deviations. *P*<0.05 indicates a significant statistical difference.

## Results

### Demographic information

Among the 3097 participants in this study, 1595 were male (approximately 51.50%). The average age of the sample is 54.46 years (SD 14.38), and the average educational level is 9.49 years (SD 4.61). The specific distribution can be seen in **Table 1**.

**Table 1.** Demographics distribution of 3097 participants.

| Age group, y | Education level |  |  |  |  |  |  |  | Total by age |  |
| --- | --- | --- | --- | --- | --- | --- | --- | --- | --- | --- |
|  | Illiteracy |  | Primary school |  | Middle school |  | High school and above |  |  |  |
|  | M | F | M | F | M | F | M | F | M | F |
| <40 | - | - | - | - | 29 | 24 | 163 | 196 | 192 | 220 |
| 40-49 | 5 | 19 | 36 | 60 | 106 | 118 | 157 | 140 | 304 | 337 |
| 50-59 | 5 | 24 | 94 | 143 | 200 | 148 | 194 | 163 | 493 | 478 |
| 60-69 | 18 | 41 | 90 | 60 | 87 | 59 | 133 | 82 | 328 | 242 |
| 70-79 | 19 | 49 | 65 | 52 | 50 | 24 | 57 | 38 | 191 | 163 |
| ≥80 | 10 | 18 | 41 | 20 | 19 | 11 | 17 | 13 | 87 | 62 |
| Total by education | 57 | 151 | 326 | 335 | 491 | 384 | 721 | 632 | 1595 | 1502 |

M: Male; F: Female; y: years.

### Demographic influences on the MoCA score

Multivariate linear regression analysis was used to assess the impact of demographic information on the total score of MoCA and its sub-cognitive domains. The regression model incorporating age and years of education demonstrated the highest predictive capability for the MoCA total score(adjusted R^2^ =0.468, F=905.778, *p*<0.05), explaining 46.8% of the variance. In the regression analysis, increasing age (*p*<0.05), less education (*p*<0.05) were associated with a lower MoCA total score, as shown in **Table 2** and **Figure 1**. It was also found that age, education, and gender predicted the scores of each sub cognitive domain to varying degrees. The detailed results of multiple linear regression analysis can be seen in **Table 2**.

**Table 2.**
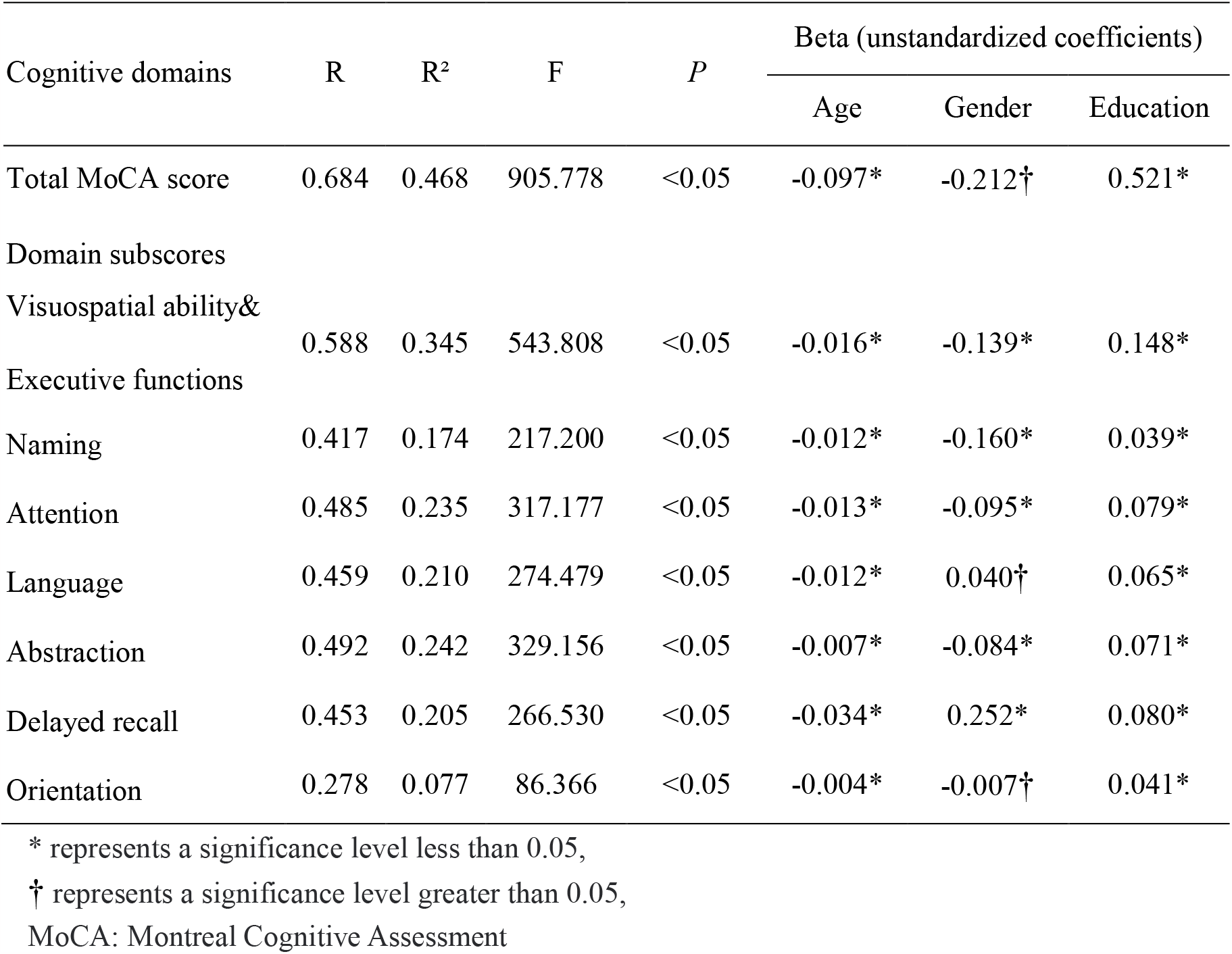
Multiple linear regression and beta unstandardised coefficients for the total MoCA score and for the cognitive domain subscores.

**Figure 1.**
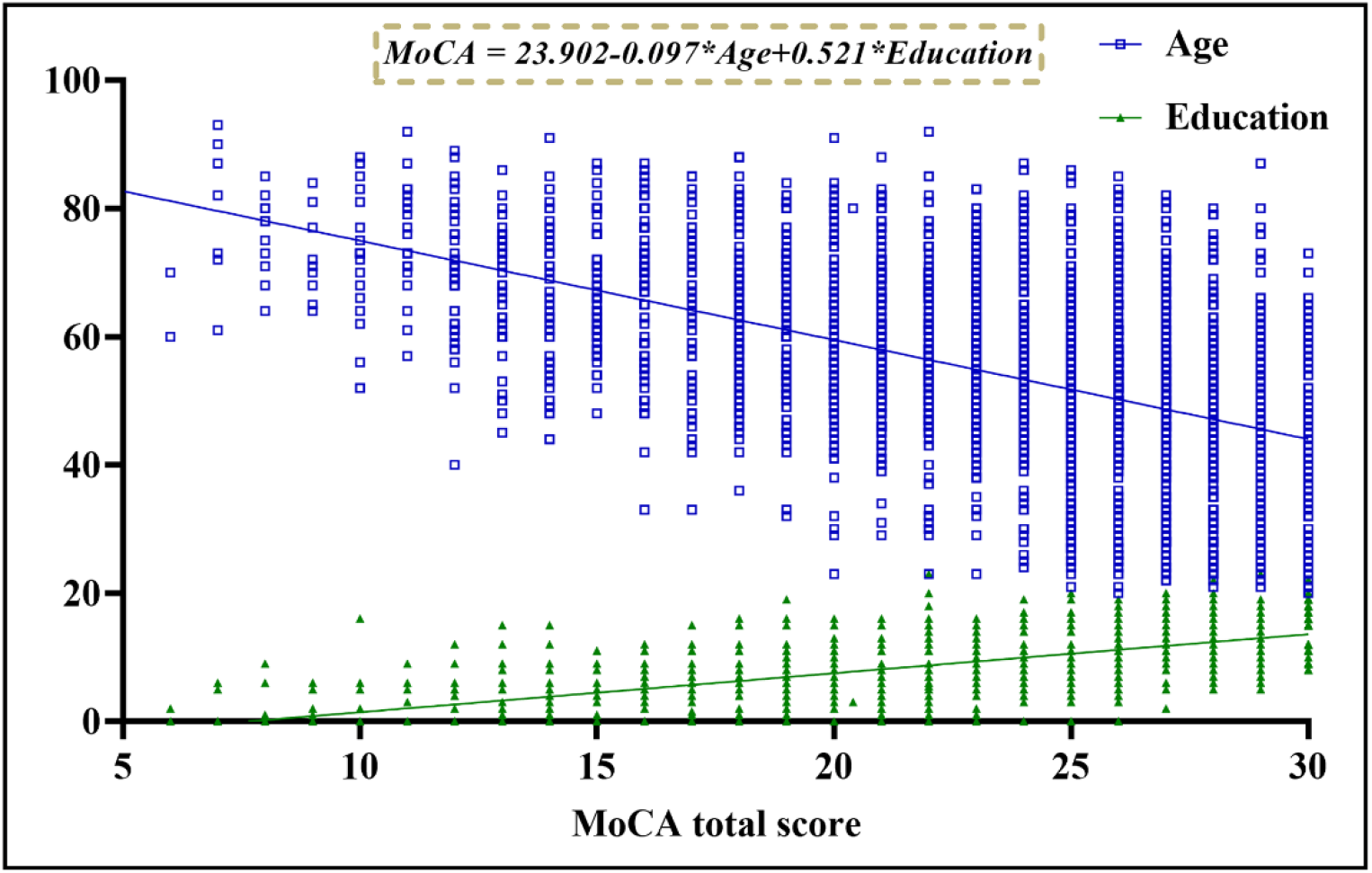
Association of the MoCA total score with age and education. The blue square represents age, and the green triangle represents education. The total score of MoCA is significantly correlated with age and education, and the specific regression equation is (MoCA=23.902-0.097 * Age+0.521 * Education). MoCA: Montreal Cognitive Assessment

### MoCA total score

Normative data stratified by age and education were derived. The mean MoCA score of the participants was 23.25 (SD 4.82) and was higher among the youngest participants and those with the highest education. Due to the popularization of education, no participants under the age of 40 with an education level of illiterate or elementary school were recruited. Normative data stratified by age and education can be seen in **Table 3**. As education level decreases, MoCA total score decrease faster with age, as shown in **Figure 2**. We provide data for calculating cut-off scores ≤ 1, ≤ 1.5, and ≤ 2 SD below the mean score in **Table 4** and **Figure 3**.

**Table 3.**
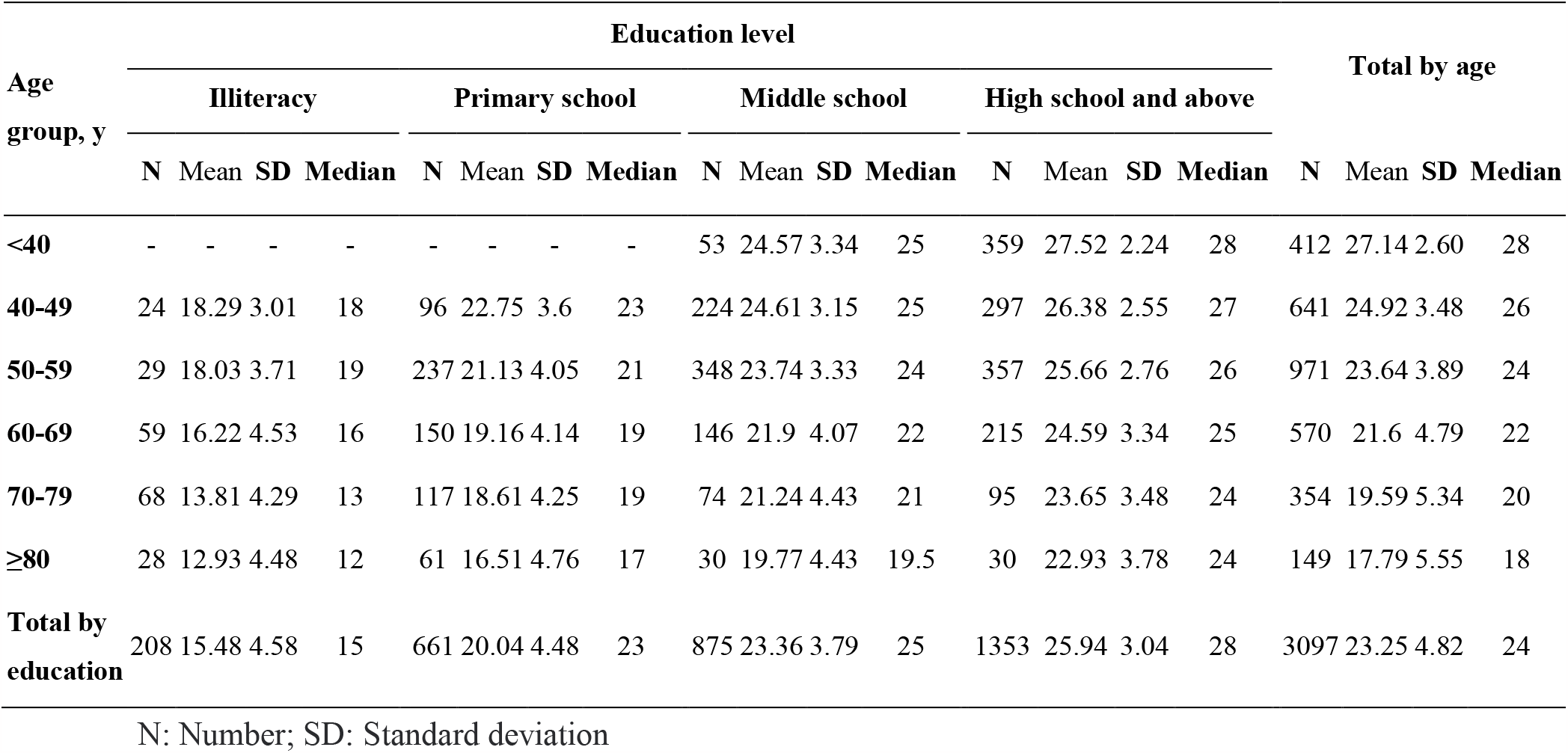
MoCA total score by age and education level.

**Table 4.** Cut-off scores by age and education level.

| Age group, y | SD below mean | Education level |  |  |  |
| --- | --- | --- | --- | --- | --- |
|  |  | Illiteracy | Primary school | Middle school | High school and above |
| <40 | ≤1 | - | - | 21 | 25 |
|  | ≤1.5 | - | - | 20 | 24 |
|  | ≤2 | - | - | 18 | 23 |
| 40-49 | ≤1 | 15 | 19 | 21 | 24 |
|  | ≤1.5 | 14 | 17 | 20 | 23 |
|  | ≤2 | 12 | 16 | 18 | 21 |
| 50-59 | ≤1 | 14 | 17 | 20 | 23 |
|  | ≤1.5 | 12 | 15 | 19 | 22 |
|  | ≤2 | 11 | 13 | 17 | 20 |
| 60-69 | ≤1 | 12 | 15 | 18 | 21 |
|  | ≤1.5 | 9 | 13 | 16 | 20 |
|  | ≤2 | 7 | 11 | 14 | 18 |
| 70-79 | ≤1 | 10 | 14 | 17 | 20 |
|  | ≤1.5 | 7 | 12 | 15 | 18 |
|  | ≤2 | 5 | 10 | 12 | 17 |
| ≥80 | ≤1 | 8 | 12 | 15 | 19 |
|  | ≤1.5 | 6 | 9 | 13 | 17 |
|  | ≤2 | 4 | 7 | 11 | 15 |

y: Years; SD: Standard deviation

**Figure 2.**
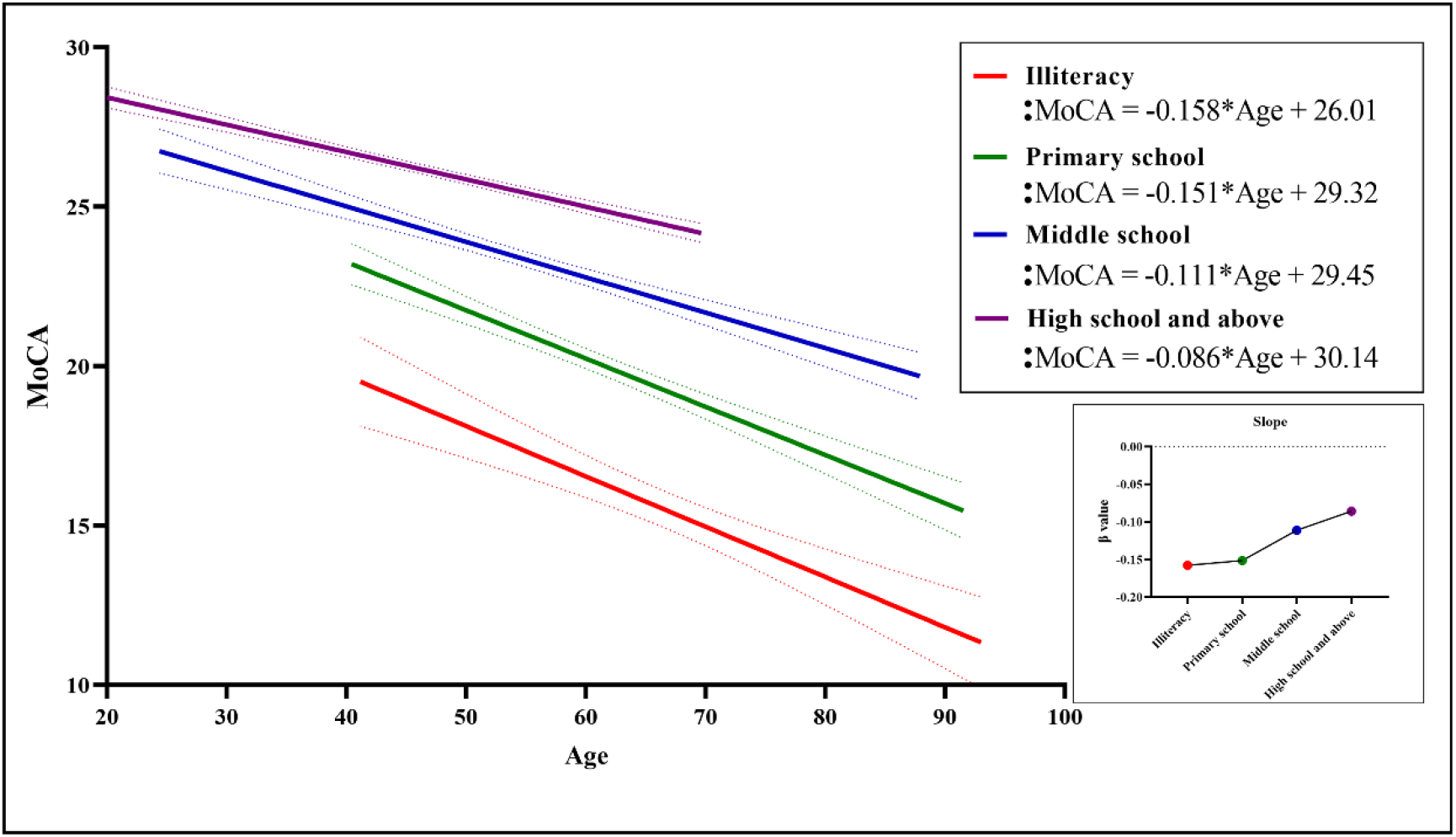
Total MoCA scores based on age for different levels of education. A negative correlation between MoCA scores and age can be observed in different educational levels, and cognitive decline significantly slows down with increasing education level. MoCA: Montreal Cognitive Assessment

**Figure 3.**
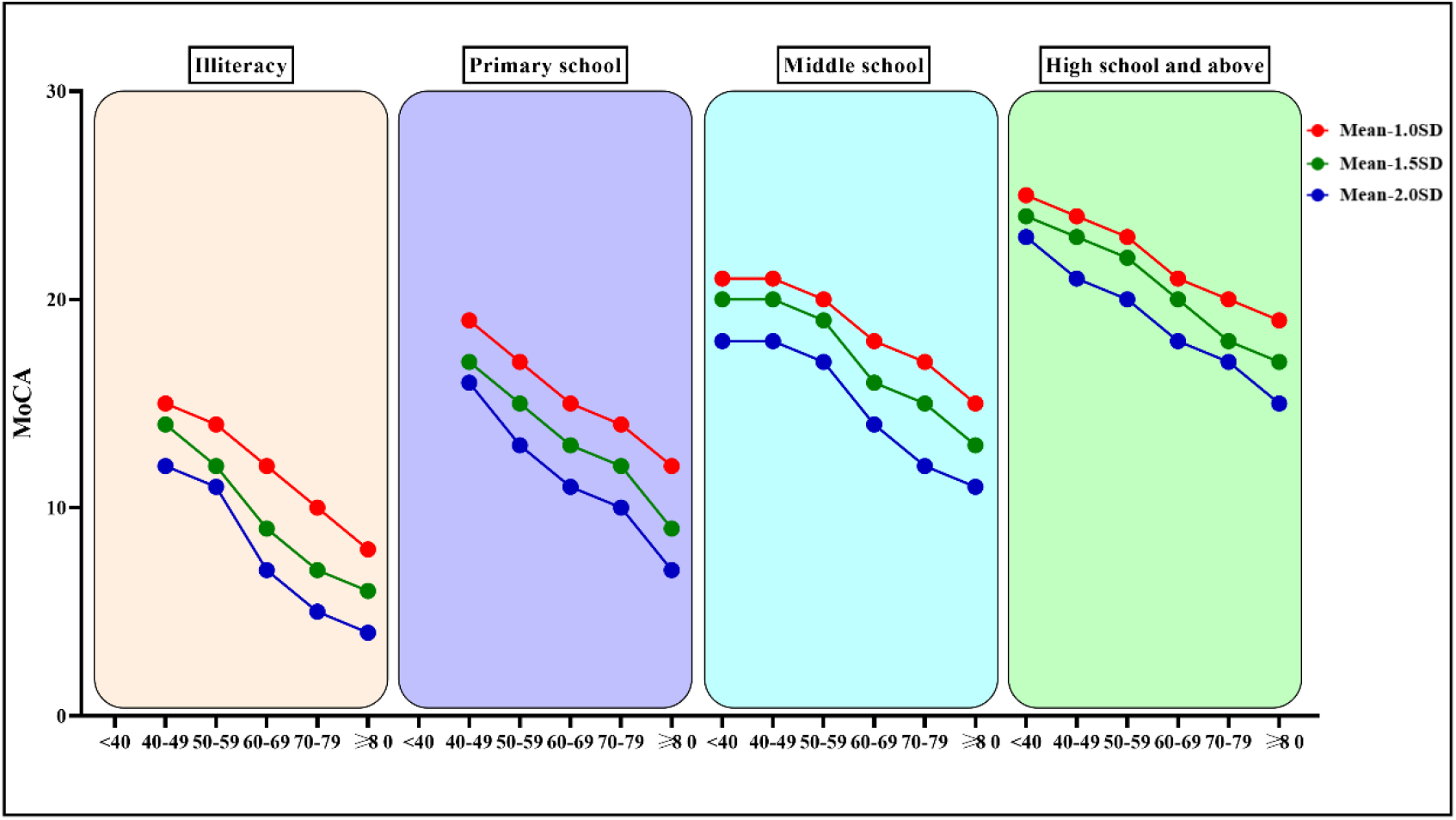
Cut-off scores based on age for different levels of education. The cut-off scores show a decreasing trend with age in all levels of education.

## Discussion

This study presents normative MoCA data stratified by age and education for individuals in Mainland China. The data were collected from a large, population-based sample, including healthy adults from 22 provinces, autonomous regions, and municipalities directly under the central government, thus providing a good representation of the Chinese population to a certain extent.

Over an extended period, the MoCA scale has been used by multiple countries and cultural systems to assess the cognitive impairment of the population, especially due to the screening of MCI. Throughout this duration, the MoCA scale has undergone translation into various iterations and gained widespread adoption. Currently, there are Beijing version 7.1, Cantonese version 7.1, Changsha version 7.1, Hong Kong version 7.1, and so on for Chinese. For this study, the latest revised Mandarin-8.1 version was employed. This iteration of the scale has been translated and revised from the original English 8.1 version. In comparison to preceding renditions, it more accurately aligns with the linguistic attributes and cultural norms of the Chinese population, rendering it better suited for the Chinese context. Consequently, it has the potential to yield more rational, consistent, and dependable test scores.

In earlier work,Nasreddine et al. previously set a score of 26 as the cut-off value for cognitive impairment, a benchmark that has garnered widespread recognition among researchers for an extended period of time. This study further revealed that the previously established cut-off value of 26 points was not suitable for the Chinese population. Among the 3,097 participants in this study, merely 1,204 (38.88%) achieved a score equal to or higher than the conventional cut-off points of 26, as proposed by Nasreddine et al..^[8]^ This outcome did not surpass our anticipated psychological expectations. Moreover, similar observations have been documented in studies focused on standardized MoCA data across diverse countries. These investigations have revealed that relatively few participants attain a score of 26, and a substantial proportion of participants exhibit younger ages or possess higher education levels.^[13, 21-23, 34]^ Moreover, a recent study demonstrated that although the critical value of 26 points exhibits high sensitivity in detecting dementia, its specificity is comparatively suboptimal.^[13]^ Therefore, it is even more important for the Chinese population to develop more realistic MoCA standardized values. It is important to highlight that the study conducted by Nasreddine et al. differs significantly from our research in terms of research design and objectives. Their study primarily aimed to ascertain the most effective score for distinguishing between healthy volunteers, individuals with mild cognitive impairment, and dementia patients, with a relatively higher educational level among the subjects.^[35]^ In contrast, our research focused on generating normative MoCA data applicable to diverse age groups and education levels among the Chinese population. As illustrated in Table 1, this study encompasses potential reasons for the substantial variance observed in the 26-point cut-off when compared to our research findings.

It is widely recognized that demographic factors such as age and educational attainment exert a substantial influence on cognitive function. In this study, multiple linear regression analysis demonstrated that MoCA scores are predominantly influenced by years of education and age, while no statistically significant relationship was observed with gender. These results are consistent with previous studies on MoCA, which have sparked considerable debate over the potential impact of gender on the total score of MoCA.^[14-16, 18, 21, 22, 34, 36]^ Age and education accounted for 46.8% of the variance in MoCA raw scores, a proportion that is notably higher than the normative data reported in recent years in some other countries.^[23, 24, 31, 32]^ This could be attributed to our substantial sample size and comprehensive representation of various age groups and education levels.[24] This study aligns with the findings of previous research, as it indicates that younger age and higher education levels are associated with higher MoCA scores.^[14, 15, 17, 19-21, 23, 33, 37, 38]^ Hence, when assessing cognitive impairment using MoCA, it is imperative to take into comprehensive account the subjects’ age and educational background. Across the four distinct educational strata examined in this study, a discernible pattern emerged wherein MoCA scores exhibited a reduction in tandem with advancing age. Moreover, our investigation revealed a noteworthy observation: individuals possessing higher educational qualifications demonstrated comparatively attenuated cognitive decline in relation to age. This overarching trend is graphically depicted in Figure 2. This aligns coherently with prior research outcomes that have similarly underscored the positive correlation between higher education levels and superior cognitive function. Essentially, education serves as a protective factor for cognitive function.^[39-42]^

It must be mentioned that this study also has some limitations. Firstly, despite the wide geographical distribution of participants, there exist substantial variations in the number of enrolled individuals across different regions. Secondly, even if there are no obvious complaints of cognitive impairment, we did not screen for too many potential factors that may affect cognitive function (excluding stroke) and did not use strict cognitive assessments to evaluate participants’ cognitive function. Therefore, it remains possible that latent cognitive impairments among the subjects cannot be entirely ruled out. Thirdly, we did not employ MMSE to further evaluate the consistency of the revised MoCA, a choice that also partially mitigated the potential bias stemming from the learning effects associated with administering analogous tests.

Conversely, it is essential to highlight certain advantages of this study. The MoCA scale employed here has undergone localization and revision to align with the characteristics and language habits of the Chinese population. The participants’ representativeness is relatively high, and the study encompasses a large sample size. Therefore, the study results are more indicative of the actual conditions prevalent among the Chinese population.

## Conclusion

This study provides normative values for MoCA that are more suitable for the Chinese population. Our research findings further confirm the contribution of age and years of education to the total MoCA score, and further indicate that the cut-off value of 26 points is not suitable for the Chinese population. And we provide boundary values based on different ages and years of education.

## Acknowledgments

The authors appreciate the contributions from all the investigators, participants.

## Author contributions

Qiang Wei: concept, literature search, statistical analysis, manuscript preparation, Baogen Du: literature search, data acquisition, statistical analysis, manuscript editing, Yuanyuan Liu: data acquisition, data analysis, Shanshan Cao: data analysis, statistical analysis, Shanshan Yin: data acquisition, data analysis, Ying Zhang: data acquisition, data analysis, Tongjian Bai: literature search, statistical analysis, Xingqi Wu: literature search, data analysis, Yanghua Tian: concept, manuscript review, Panpan Hu: concept, design, manuscript review, Kai Wang: concept, design, manuscript review.

## Data availability

All datasets generated in this study are included in the manuscript. The data supporting the findings of this study are available on request from the corresponding author.

